# Post-pandemic memory T-cell response to SARS-CoV-2 is durable, broadly targeted and cross-reactive to hypermutated BA.2.86

**DOI:** 10.1101/2023.10.28.23297714

**Authors:** Rofhiwa Nesamari, Millicent A. Omondi, Maxine A. Höft, Amkele Ngomti, Richard Baguma, Anathi A. Nkayi, Asiphe S. Besethi, Siyabulela F. J. Magugu, Paballo Mosala, Avril Walters, Gesina M. Clark, Mathilda Mennen, Sango Skelem, Marguerite Adriaanse, Alba Grifoni, Alessandro Sette, Roanne S. Keeton, Ntobeko A.B. Ntusi, Catherine Riou, Wendy A. Burgers

## Abstract

The COVID-19 post-pandemic period is characterised by infection waves of uncertain size (due to low rates of SARS-CoV-2 testing and notification), as well as limited uptake or global access to updated variant vaccines. Ongoing SARS-CoV-2 evolution has given rise to recombinant Omicron lineages that dominate globally (XBB.1), as well as the emergence of hypermutated variants (BA.2.86). In this context, durable and cross-reactive T-cell immune memory is critical for continued protection against severe COVID-19. We examined T-cell responses to SARS-CoV-2 approximately 1.5 years since Omicron first emerged. We describe sustained CD4+ and CD8+ spike-specific T-cell memory responses in healthcare workers in South Africa (n=39), most of whom had received 2 doses of Ad26.CoV2.S (Johnson & Johnson/Janssen) vaccine and experienced at least one SARS-CoV-2 infection. Spike-specific T cells were highly cross-reactive with all Omicron variants tested, including BA.2.86. Abundant non-spike (nucleocapsid and membrane)-specific T cells were detectable in most participants, augmenting the total T-cell resources available for protection. The bulk of SARS-CoV-2-specific T-cell responses had an early-differentiated phenotype, explaining their persistent nature. Thus, hybrid immunity leads to the accumulation of spike and non-spike T cells evident 3.5 years after the start of the pandemic, with preserved recognition of highly mutated SARS-CoV-2 variants. Long-term T-cell immune memory is likely to provide continued protection against severe outcomes of COVID-19.

**In Brief:** Nesamari *et al.* investigate T-cell responses in the context of hybrid immunity 3.5 years after the start of the COVID-19 pandemic. They show that T-cell memory is highly durable and cross-reactive with recombinant variants XBB.1 and hypermutated BA.2.86. Abundant non-spike responses augment the overall T-cell response.

## INTRODUCTION

Sustained and cross-reactive immunological memory in the post-pandemic period is essential for continued protection from severe outcomes of COVID-19. Continued viral evolution has led to the emergence and dominance of the XBB recombinant sub-lineages of Omicron ^1^. Recently, the novel Omicron subvariant BA.2.86 was described, with up to 60 amino acid changes compared to ancestral SARS-CoV-2 in the spike protein, and over 30 changes in spike compared to the BA.2 and XBB.1.5 variants. Due to its hyper-mutated nature, BA.2.86 has been classified as a “variant under monitoring” by WHO, and as of October 26, 2023, it has been identified in 1,070 sequences from 34 countries. This is most likely an underestimate of BA.2.86 prevalence, given the current limited SARS-CoV-2 surveillance effort. Several recent studies have evaluated the neutralization sensitivity of BA.2.86 ^2–5^. As anticipated, BA.2.86 shows extensive immune evasion relative to ancestral SARS-CoV-2 in sera collected prior to the Omicron wave ^2^; although in BA.1-infected individuals, the degree of neutralization of BA.2.86 was similar to that of XBB lineages currently dominating globally. However, the ability of spike-specific T cells to cross-recognize BA.2.86 spike has not yet been investigated. While it has been demonstrated that spike T-cell responses generated upon natural infection and vaccination against the ancestral SARS-CoV-2 spike are highly cross-reactive against Omicron BA.1 ^6–9^, it is important to determine whether the extensive mutations in BA.2.86 spike could hinder its recognition by spike memory T-cell responses in individuals who have been infected and/or vaccinated during the course of the COVID-19 pandemic.

It is now clearly established that the SARS-CoV-2-specific antibody response (generated upon infection or vaccination) wanes relatively quickly ^10^ and shows reduced neutralization activity against each new variant of concern (VOC) that dominates circulation, resulting in sub-optimal protection against SARS-CoV-2 infection. In contrast, memory T-cell responses to SARS-CoV-2 can persist for up to a year following exposure to the SARS-CoV-2 spike protein ^11–14^ and maintain robust cross-reactivity against VOCs. As we find ourselves three and a half years into the COVID-19 pandemic, where infection waves are smaller and booster vaccination is limited in most parts of the world due to restricted eligibility or availability, it is critical that the long-term durability of SARS-CoV-2-specific T-cell responses is monitored.

In this study, we included 39 healthcare workers, with a documented history of SARS-CoV-2 infection and vaccination, to determine whether their prevailing spike-specific memory T-cell responses in mid-late 2023, could cross-recognize the BA.2.86 sub-lineage. T-cell cross-reactivity was assessed in both in Omicron-infected and -uninfected participants. In parallel, paired samples obtained two years apart were used to explore SARS-CoV-2-specific T-cell durability, including assessing the contribution of non-spike T cell responses to overall T-cell immunity.

## RESULTS

### Cross-reactivity of spike-specific T-cell response to Omicron variants

We measured T-cell responses to spike in blood samples (n=39) collected between July and September 2023. At this timepoint (T2 in **Table 1** and **Fig. S1**), 28.2% (11/39) of the participants had received one dose of Ad26.COV2.S vaccine, 56.4% (22/39) two vaccine doses and 15.4% (6/39) three vaccine doses. The median time since last vaccination was ∼21 months (IQR: 20.2-24.4). Twenty-two participants (56.4%) had a documented SARS-CoV-2 infection prior to the onset of the Omicron wave, and all experienced a breakthrough infection during the Omicron wave, at a median time of 19.4 months (IQR: 17.8-19.9) before sample collection. We measured cytokine production (IFN-γ, IL-2 and TNF-α) by intracellular cytokine staining in response to peptide pools covering the full ancestral, BA.1, XBB.1 or BA.2.86 spike protein (**Fig. 1A**). **Fig. 1B** shows the frequency of memory CD4+ T-cells to each SARS-CoV-2 spike tested. Notably, the majority of participants (94.9%) still exhibited a robust ancestral spike-specific CD4+ T-cell response (median: 0.031%, IQR: 0.018-0.059) one and a half years after their last known SARS-CoV-2 infection. When comparing reactivity to ancestral, BA.1, XBB.1 and BA.2.86 spike, we observed no significant difference in the frequency of spike-specific CD4+ T-cells between SARS-CoV-2 variants (**Fig. 1B**). For each Omicron sub-lineage, the fold change in the frequency of spike-specific CD4+ T-cell responses, relative to the ancestral spike, was calculated (**Fig. 1C**). Overall, the spike-specific CD4+ T-cell response was highly preserved (≥90%) against all Omicron variants tested, including the hyper-mutated BA.2.86. We also assessed the cross-reactivity of spike-specific CD8+ T-cell responses. In contrast to the CD4 compartment, the proportion of CD8 responders to ancestral spike was strikingly lower (∼40%), and this was consistent amongst all three Omicron sub-lineages tested. While the median magnitude of spike-specific CD8+ T-cell responses was similar across all variants (**Fig. 1D**), the fold change profile within individual participants was variable. A fraction of participants who did not have a detectable CD8 response to ancestral spike (5/24, 20.8%) gained a CD8 response to one or more Omicron sub-lineages, likely reflecting of *de novo* generation of a CD8 response to their Omicron breakthrough infection. In participants who had a detectable CD8+ T-cell response to ancestral spike, at least 50% of the CD8+ T-cell response was preserved against Omicron sub-lineages in most participants (10/15 for BA.1 and BA.2.86 and 12/15 for XBB.1), while a small fraction of individuals exhibited a reduction (>50%) or loss in T-cell reactivity to Omicron spike (5/15 for BA.1 and BA.2.86 and 3/15 for XBB.1) (**Fig. 1E**).

**Table 1:** Clinical characteristics of study participants. T1 samples were collected approximately 4-6 months prior to the Omicron BA.1 wave and T2 samples were collected 2 years later, approximately 1.5 years after the BA.1 wave (see **Fig. S1**). The majority of participants (89.7%) were vaccinated with Ad26.COV2.S. Three participants received a heterologous vaccination regimen (Ad26.COV2.S and BNT162b2) and one participant received 3 doses of the BNT162b2 vaccine. Prior infection and breakthrough infection (BTI) were determined by PCR (‘recorded infection’) or by Nucleocapsid seroconversion or a two-fold increase in Nucleocapsid-specific IgG. IQR: Interquartile range; BTI: breakthrough infection; ^a^: Ancestral SARS-CoV-2 or Beta variant infection; na: Not applicable; ^b^: PCR data available for 5/9 participants with documented infection; ^c^: PCR data available for 15/39 participants.

|  | T1 samples<br>(pre-BA.1 wave) | T2 samples<br>(post-BA.1 wave) |
| --- | --- | --- |
| n | n = 16 | n = 39 |
| Age at enrolment (median, IQR) | 51 [38-54] | 47 [31-54] |
| Gender (% female) | 87.5% | 84.2% |
| Sampling dates | July - Sept 2021 | July - Sept 2023 |
| Vaccination history |  |  |
| 1 vaccine dose (% , n) | 100% | 28.2% (n=11) |
| 2 vaccine doses (% , n) | 0% | 56.4% (n=22) |
| 3 vaccine doses (% , n) | 0% | 15.4% (n=6) |
| Months since last vaccination (median, IQR) | 5.2 [5-6] | 20.7 [20.2-24.4] |
| Infection history |  |  |
| Prior recorded infection* (% , n) | 56.2% (n=9) | 56.4% (n=22) |
| Omicron BTI (% , n) | na | 100% (n=39) |
| Months since last recorded infection (median, IQR) | 8.4 [7-13]<sup>&</sup> | 19.4 [17.8-19.9]<sup>#</sup> |
| Paired samples | n = 15 |  |
| Months between T2 and T1 samples (median, IQR) | 23.9 [23.3-24.1] |  |

**Figure 1.**
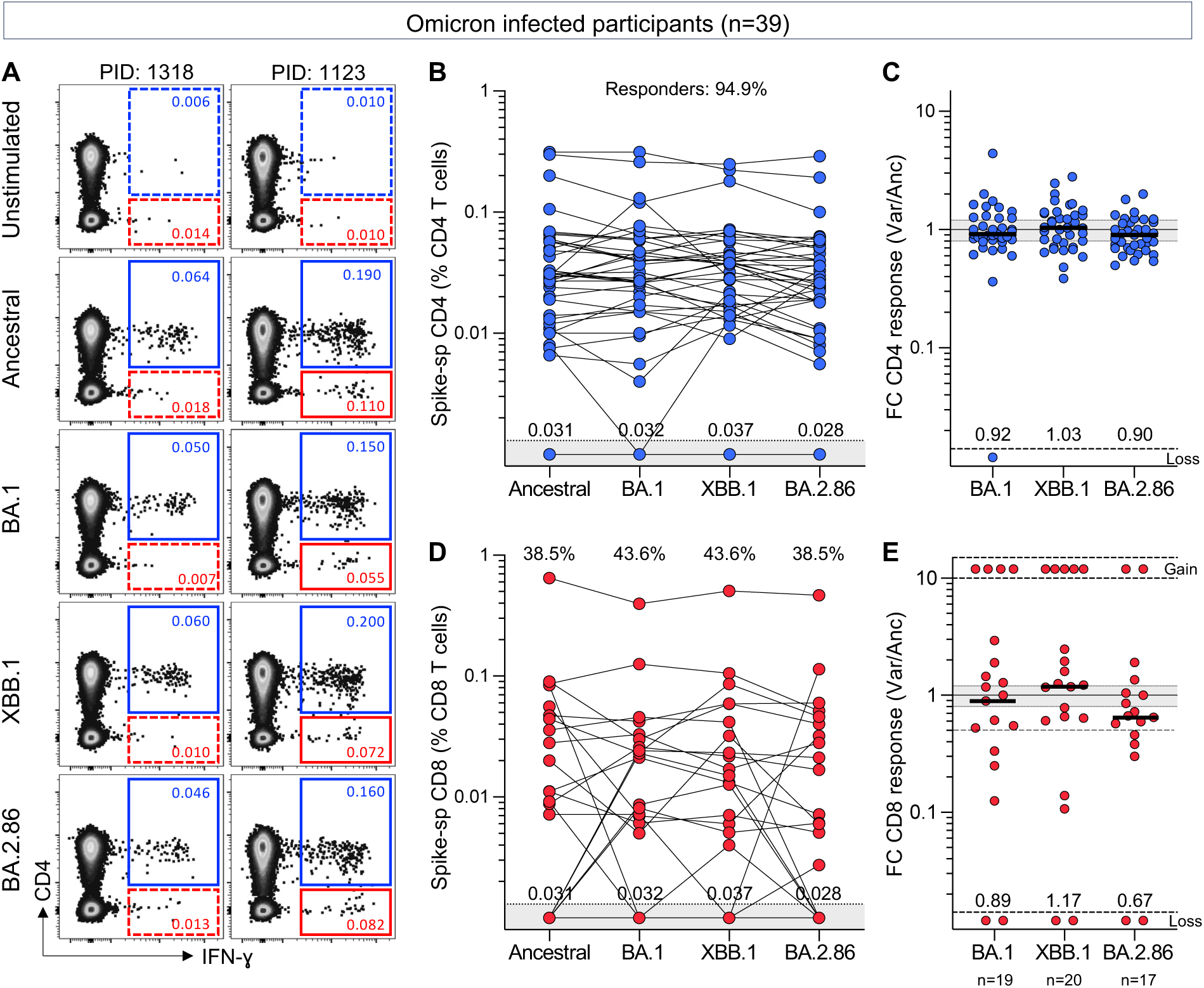
CD4+ and CD8+ T-cell responses to SARS-CoV-2 ancestral, BA.1, XBB.1 or BA.2.86 spike. **(A)** Representative examples of IFN-γ production in response to ancestral, BA.1, XBB.1 or BA.2.86 spike in two individuals. The frequency of IFN-γ+ cells is expressed as a percentage of the total CD4+ T cells (blue) or as a percentage of the total CD8+ T cells (red). **(B and D)** Frequency of spike-specific CD4+ T cells (B) and CD8+ T cells (D) producing any of the measured cytokines (IFN-γ, IL-2 or TNF-α) in 39 participants with confirmed Omicron infection. The proportion of responders is indicated at the top of the graphs. Median frequencies of spike-specific T cells in responders are indicated at the bottom of the graphs. **(C and E)** Fold change in the frequency of spike-specific CD4+ T cells (C) and CD8+ T cells (E) between ancestral and SARS-CoV-2 variants in participants with confirmed BA.1 infection. Median fold change in responders is represented by a bar and indicated at the bottom of the graphs. Gained responses are depicted on top and lost responses (“loss”) at the bottom. No significant differences were observed between variants using a Friedman test with Dunn’s multiple comparisons post-test.

Since all participants had experienced an Omicron breakthrough infection, potentially prompting the development of *de novo* T-cell responses targeting mutated epitopes of spike ^15^, we also assessed T-cell cross-reactivity at an earlier timepoint, obtained before the emergence of Omicron (see T1 in **Table 1** and **Fig. S1**). Comparable results to those obtained in post-Omicron-infected participants were found, demonstrating that spike-specific T-cell responses were highly cross-reactive with BA.1, XBB.1 and BA.2.86 (**Fig. S2**) and Omicron lineage cross-reactivity was not dependent on having been Omicron-infected.

### Nucleocapsid and membrane-specific T -response significantly contribute to memory SARS-CoV-2 adaptative immune response

To gain a more comprehensive understanding of the profile of SARS-CoV-2-specific memory T-cell responses, we defined the extent to which non-spike proteins contribute to SARS-CoV-2 immunological memory. Specifically, our focus was on the T-cell response to SARS-CoV-2 nucleocapsid and membrane proteins, as these two proteins, in addition to spike, have demonstrated the highest immunogenicity ^16–20^. T-cell responses for both spike and nucleocapsid and membrane (N&M) were available for 36 participants (**Fig. 2A**). CD4+ T-cell responses to N&M were detectable in 97.2% (35/36) of participants, with magnitudes comparable to those elicited toward spike (**Fig. 2A**, left panel). In fact, there was an association between the frequency of spike- and N&M-specific CD4+ T cells (r=0.64, p=2.3×10^-5^). In the CD8 compartment, similar proportions of responders (44.4%) were observed to spike and N&M (**Fig. 2A**, right panel). However, no association was found between spike and N&M responses (r=0.28, p=0.09), as previously reported ^21^. The overall combinations of SARS-CoV-2 CD4+ and CD8+ T-cell responses targeting spike and N&M was diverse amongst participants (**Fig. 2B**). It is noteworthy that most participants (34/36) had CD4 responses targeting both spike and N&M. In contrast, CD8 responders (23/36, 63.9%) were evenly divided amongst those who targeted both spike and N&M (9/23, 39.1%), those targeting spike exclusively (7/23, 30.4%) and those targeting N&M exclusively (7/23, 30.4%). Thus, quantifying non-spike responses increased the ability to detect CD8 responses to SARS-CoV-2 in those who were persistently spike-hyporesponsive despite multiple vaccinations. The contribution of N&M-specific T cells to SARS-CoV-2-specific memory responses is further illustrated in **Fig. 2C**, showing the profile of SARS-CoV-2 T-cell responses and clinical characteristics for each participant.

**Figure 2.**
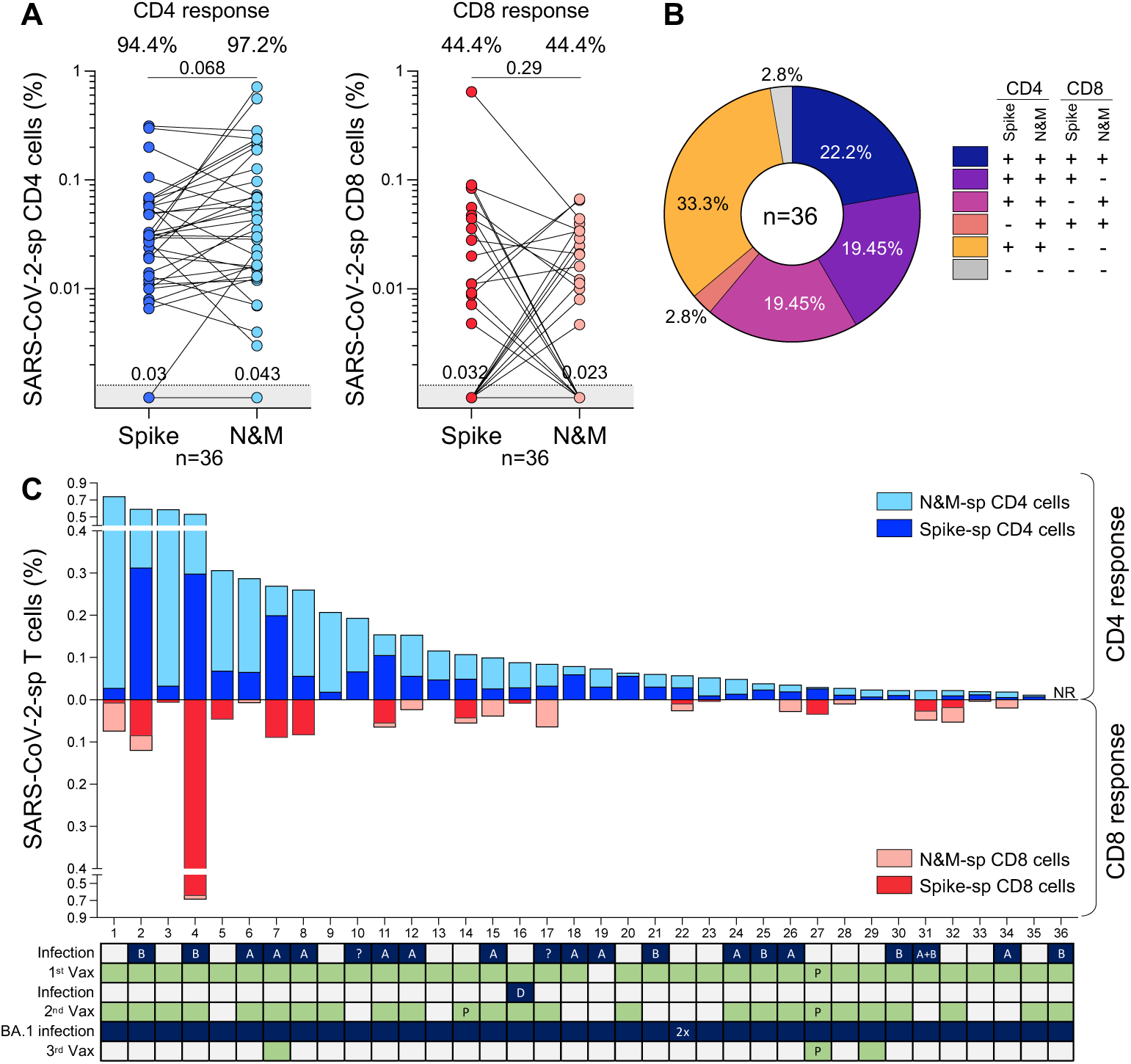
Profile of ancestral SARS-CoV-2 spike- and nucleocapsid and membrane-specific T-cell response ∼3.5 years after the start of the COVID-19 pandemic. **(A)** Frequency of spike- and nucleocapsid and membrane (N&M)-specific CD4+ (left panel) and CD8+ T cells (right panel) in 36 participants sampled between July and September 2023. Proportion of responders are indicated on top of the graphs. Medians of responders are indicated at the bottom of the graphs. Statistical comparisons were assessed using a Wilcoxon matched-pairs signed rank test. **(B)** Distribution of spike- and N&M-specific CD4+ and CD8+ T-cell responses in the study cohort. Each slice of pie represents a response pattern, as indicated in the key. **(C)** Total magnitude of spike- and N&M-specific CD4+ and CD8+ T-cell responses. The recorded SARS-CoV-2 infection and vaccination histories of each participant are indicated at the bottom of the graph. A: ancestral SARS-CoV-2 infection, B: Beta variant infection, D: Delta variant infection, ?: unknown variant infection. All vaccinations were Ad26.COV2.S vaccine, unless indicated with a “P” for Pfizer/BNT162b2.

### Durability of SARS-CoV-2 T-cell responses

Although a specific T cell-based measure of protection has yet to be defined, accumulating evidence suggests that T cells contribute to the control of SARS-CoV-2, as indicated by their associations with COVID-19 symptoms and outcomes ^22,23^. Thus, to measure T-cell maintenance, we compared the frequencies of T cells specific for spike and non-spike proteins, nucleocapsid (N) and membrane (M) in 15 paired samples. The samples were taken at two timepoints, 2 years apart: T1 (∼4-6 months prior to the Omicron BA.1 wave) and T2 (∼1.5 years after the BA.1 wave) (**Table 1**). Between the two timepoints, 60% (9/15) of the participants received a booster vaccination (median: 20.5 months before T2 sampling) and all experienced an Omicron breakthrough infection (median 19.4 months before T2 sampling). **Fig. 3A** shows the frequency of CD4+ T-cell responses to ancestral spike and N&M in these participants. No significant change was observed in the frequency of spike-specific CD4+ T cells between T1 and T2 (median: 0.036% and 0.031%, respectively), with a median fold-change variation of 0.91 (**Fig. 3B**), demonstrating a preservation of spike CD4+ T cell responses over time. Similar sustained levels of CD4+ T-cell responses were observed against BA.1, XBB.1 and BA.2.86 (**Fig. S3A and S3B**). All participants with an undetectable N&M-specific CD4+ T-cell response (n=4) had mounted a response by T2, after breakthrough infection (**Fig. 3A**). In the remaining 10 participants assessed, the preservation of N&M-specific CD4+ T cells varied, with a median fold change of 0.57 (ranging from 0.28 to 1.54) (**Fig. 3B**).

**Figure 3.**
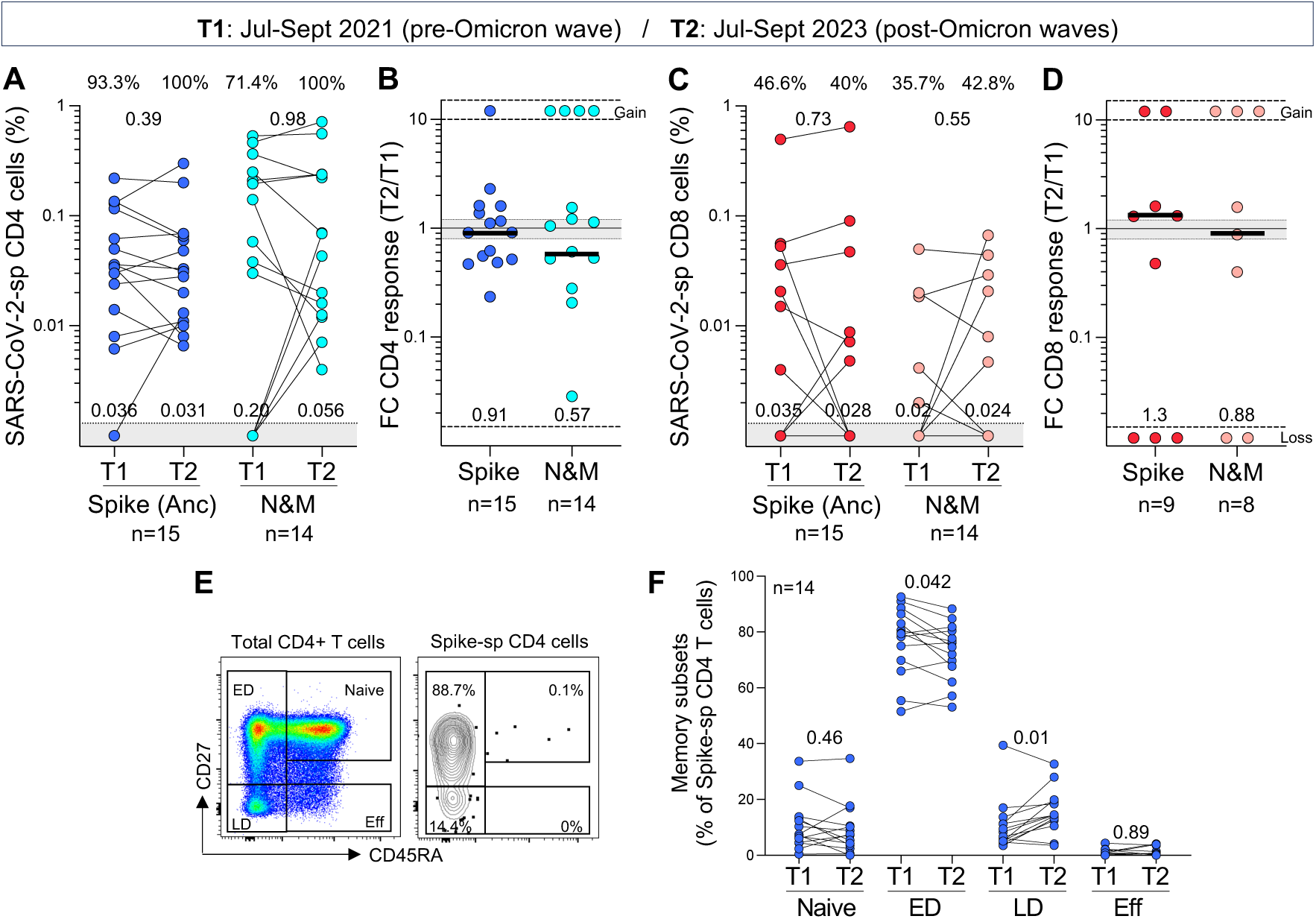
Longitudinal assessment of the maintenance and memory profile of ancestral SARS-CoV-2-specific T-cell responses over 2 years. **(A and C)** Frequency of ancestral spike- and nucleocapsid and membrane (N&M)-specific CD4+ (A) and CD8+ T cells (C) in paired samples (n=15 for Spike and n=14 for N&M). T1 and T2 samples were collected between July and September 2021 and July and September 2023, respectively. Medians of responders are indicated at the bottom of the graphs. Statistical comparisons were assessed using a Wilcoxon matched-pairs signed rank test. **(B and D)** Fold change in the frequency of SARS-CoV-2-specific CD4+ (B) and CD8+ T cells **(D)** between T2 and T1 in responders. Bars represent medians, and median fold change is indicated at the bottom of each graph. Gained responses are depicted on top and lost responses at the bottom. **(E)** Representative flow plots of the memory differentiation profile of total CD4+ T cells and ancestral spike-specific CD4+ T cells. Naïve: CD45RA+CD27+, ED (early differentiated cells): CD45RA-CD27+, LD (late differentiated cells): CD45RA-CD27- and Eff (Effector cells): CD45RA+CD27-. **(F)** Comparison of the memory profile of spike-specific CD4+ T cells (n=14) at T1 and T2. To define the memory phenotype of spike-specific T cells, a cut-off of 30 events was used. Statistical comparisons were assessed using a Wilcoxon matched-pairs signed rank test.

Within the CD8 compartment, only a small number of the paired participants exhibited detectable spike- or N&M-specific CD8+ responses (**Fig. 3C**). The evolution of these CD8+ T-cell responses from T1 to T2 was highly variable amongst participants, showing newly acquired, sustained, or lost responses (**Fig. 3D**). Similar patterns were observed for CD8+ T-cell responses against BA.1, XBB.1 or BA.2.86 (**Fig. S3C and S3D**).

Lastly, we defined the memory profile of spike-specific T cells. Using the differentiation markers CD45RA and CD27, we identified four memory subsets, namely naïve (CD45RA+CD27+), early differentiated (ED: CD45RA-CD27+), late differentiated (LD: CD45RA-CD27-) and effector (Eff: CD45RA+CD27-) (**Fig. 3E**). Ancestral spike-specific CD4+ T cells exhibited primarily an ED memory profile at T1 (**Fig. 3F**). At T2, there was a marginal decrease in the proportion of ED spike-specific CD4+ T cells (median 75.4% vs 79.5% at T1, p=0.042), which was counterbalanced by an increase in cells exhibiting a late differentiated profile (p=0.01). Of note, the memory profile of CD4+ T cells recognising BA.1, XBB.1 and BA.2.86 spike was similar to that observed for ancestral spike, characterized by a predominance of ED memory cells (**Fig. S3E**). Due to the limited number of paired samples with a spike-specific CD8+ T-cell response, we could not reliably compare the memory phenotype of these cells at the two timepoints. However, the memory profile of SARS-CoV-2-specific CD8+T cells was defined from samples collected at T2 (**Fig. S3F**). Unlike the CD4 response, spike-specific CD8+ T cells exhibited a more diverse memory profile within each participant, consisting of a median of ∼40% of ED cells, ∼20% of LD cells and ∼20% of effector cells.

## DISCUSSION

In this study we observed robust circulating memory T cells, critical components of the antiviral T-cell response, persisting in healthcare workers in mid-late 2023, >1.5 years after the global Omicron wave, with most individuals receiving their last vaccination prior to Omicron breakthrough infection. All participants experienced mild primary or breakthrough infections. Immunological memory has previously been reported up to 1 year after vaccination or infection, regardless of the severity of disease ^24^ and to our knowledge these data represent the most recent T-cell response measurements reported from the post-pandemic period. Most parts of the world now experience ongoing viral circulation ^1^. Thus, maintenance could be related to recurrent exposure to SARS-CoV-2 in some individuals, expanding the T-cell memory pool, or simply highly durable memory responses persisting from prior vaccination and infection. Phenotypic analysis revealed that SARS-CoV-2-specific T cells exhibit predominantly an early differentiated memory phenotype, consistent with several studies ^25,26^. It is noteworthy that SARS-CoV-1 responses were detectable up to 17 years after infection ^27,28^. Together, these data suggest a high capacity for the T-cell response to persist long term and provide potent recall responses upon SARS-CoV-2 re-exposure, even in the absence of booster vaccination or viral exposure.

We also demonstrate that T-cell responses are able to effectively cross-recognize SARS-CoV-2 variants, including XBB.1 whose sub-lineages currently dominate globally ^1^. Moreover, cross-reactivity was preserved to the highly mutated BA.2.86, with >60 mutations in spike compared to the ancestral virus, in samples collected prior to the emergence of BA.2.86. This retention of T-cell reactivity across variants is consistent with many published studies ^6–9,29^, emphasizing the potency of T-cell responses against a backdrop of diminishing neutralizing antibody responses with successively more highly evolved variants ^2–5^. Importantly, we observed that Omicron sub-lineage cross-reactivity was readily detectable even before Omicron infection, suggesting that the T-cell response to conserved spike epitopes, included in the first-generation vaccines, may provide adequate cross-protection. This is reassuring, given that the availability of updated booster vaccines based on XBB.1.5 in late 2023 ^30^ is largely restricted to high income countries. The stable preservation of CD4 responses across variants suggests that most targeted epitopes are located in non-variable parts of spike or that mutations do not affect epitope recognition ^31^. In contrast, as previously reported ^6,9^, the preservation of CD8+ T-cell responses to variants is more heterogenous. Our results emphasize that while variant mutations may lead to the occasional loss of epitope cross-recognition, they could also result in the creation of new immunogenic epitopes after breakthrough infection. Together, these data demonstrate that highly resilient and adaptable T-cell responses are present in most individuals in the post-pandemic period.

An important consideration of hybrid immunity is that infection delivers additional T cell targets from the SARS-CoV-2 proteome. We showed that non-spike T-cell responses constituted a sizable portion of the overall SARS-CoV-2 response, expanding the breadth of the response from vaccination. This was particularly striking for CD8 responses, with a third of responders targeting nucleocapsid and membrane in the absence of a CD8 spike response, consistent with epitope repertoires highly dependent on the HLA background of the individual ^24,31^. Unlike the neutralization response, T cells targeting spike or non-spike antigens have the potential to clear infected cells and limit viral replication. Since non-spike proteins are not under relentless selective pressure from neutralizing antibodies, accumulation of mutations is limited, ensuring a high degree of T-cell cross-reactivity to emerging variants ^32^. For these reasons, conserved non-spike Sarbecovirus epitopes are being included in pan-coronavirus vaccines in development ^33^. Overall, we report durable, broad and highly cross-reactive post-pandemic T-cell responses in healthcare workers who were vaccinated and infected with SARS-CoV-2. The results of this study demonstrate that long-term immunological T-cell memory persists in the background of heterogenous exposure history, and withstands continued and extensive viral variation, providing immune resources for protection from severe outcomes of SARS-CoV-2 infection.

### Limitations of the study

Our study had several limitations. We may have underestimated the number of additional infections in subsequent smaller waves that have followed the initial Omicron BA.1 wave. Testing is no longer free or easily accessible, SARS-CoV-2 has ceased to be a notifiable disease and asymptomatic infections are more likely, given substantial population immunity ^34^. Memory responses after recorded Omicron breakthrough infections may thus have been boosted with further exposures, influencing durability, magnitude and cross-reactivity. Furthermore, while structural proteins are the dominant T-cell targets ^35^, we did not measure responses to non-structural components of the viral proteome ^16^ and thus may have underestimated the total T-cell response. Lastly, we were restricted to measuring T cells in circulation, but infection can lead to sequestration of memory T cells to the respiratory tract, that may offer protection at the sites of infection ^36,37^. Further studies are needed to examine the durability of these tissue resident memory T cells.

## Data Availability

All data reported in this paper will be shared by the lead contacts upon request. This paper does not report original code. Any additional information required to reanalyze the data reported in this paper is available from the lead contacts upon request.

## ACKNOWLEDGEMENTS

We thank the study participants and the clinical staff and personnel at Groote Schuur Hospital, Cape Town for their dedication. Research reported in this publication was supported by the Wellcome Trust (226137/Z/22/Z), the South African Medical Research Council (SA-MRC) with funds received from the South African Department of Science and Innovation (DSI), the Bill and Melinda Gates Foundation (INV-046743), the Poliomyelitis Research Foundation (21/65) and the Wellcome Centre for Infectious Diseases Research in Africa (CIDRI-Africa), which is supported by core funding from the Wellcome Trust (203135/Z/16/Z and 222754). W.A.B. and C.R. are supported by the EDCTP2 program of the European Union’s Horizon 2020 program (TMA2016SF-1535-CaTCH-22 to W.A.B and TMA2017SF-1951-TB-SPEC to C.R.). R.N. is supported by the Harry Crossley Foundation. N.A.B.N. acknowledges funding from the SA-MRC, MRC UK, NRF, and the Lily and Ernst Hausmann Trust. This project has been funded in part with Federal funds from the National Institute of Allergy and Infectious Diseases, National Institutes of Health, Department of Health and Human Services, under Contract No. 75N93021C00016 to A.G. and Contract No. 75N93019C00065 to A.S. For the purposes of open access, the authors have applied a CC-BY public copyright license to any author-accepted version arising from this submission.

## AUTHOR CONTRIBUTIONS

W.A.B., C.R., and R.N. conceived the study, designed the experiments, analyzed the data, and wrote the manuscript. R.N., R.K. M.A.O., M.A.H., A.N., R.B., S.F.G.M., A.S.B., A.A.N., P.M., G.M.C., and A.W. performed the experimental work. N.A.B.N. established and led the HCW cohort. M.M., S.S., and M.A. recruited participants, managed the HCW cohort and contributed to clinical samples. A.G. and A.S. designed and provided peptide pools. All authors reviewed and edited the manuscript.

## STAR★METHODS

### RESOURCE AVAILABILITY

#### Lead contact

Further information and requests for resources and reagents should be directed to and will be fulfilled by the lead contact: Wendy Burgers.

#### Materials availability

Materials will be made available from the lead contact with a completed Materials Transfer Agreement.

### EXPERIMENTAL MODEL AND SUBJECT DETAILS

#### Human Subjects

Participants included in this study (n=40) were selected from a longitudinal study of healthcare workers (HCW) enrolled from Groote Schuur Hospital (Cape Town, Western Cape, South Africa) ^38,39^. Participants were selected based on the availability of peripheral blood mononuclear cells (PBMC) in our biorepository. We used samples collected at two timepoints: 1) Timepoint 1 samples (T1, n=16) were collected between July and September 2021 (4-6 months prior to the Omicron BA.1 wave). At this timepoint, 9 out of 16 (56.2%) had a recorded SARS-CoV-2 infection and all participants received one dose of the Ad26.COV2.S vaccine (Johnson and Johnson/Janssen) 5.2 months [IQR: 5-6] prior to blood collection; 2) Timepoint 2 samples (T2, n=39) were collected between July and September 2023, approximately 1.5 years after the BA.1 wave. At this timepoint, 28.2% (11/39) of the participants have received one Ad26.COV2.S vaccine dose, 56.4% (22/39) two vaccine doses and 15.4% (6/39) three vaccine doses. The majority of participants (89.7%) were vaccinated with Ad26.COV2.S. Three participants received a heterologous vaccination regimen (Ad26.COV2.S and BNT162b2) and one participant received 3 doses of the BNT162b2 vaccine. The median time since last vaccination was ∼21 months (IQR: 20.2-24.4). Twenty-two participants (56.4%) had a documented SARS-CoV-2 infection, prior to the onset of the Omicron wave; and all experienced a breakthrough infection during the Omicron wave, at a median time of 19.4 month (IQR: 17.8-19.9) before sample collection. The landscape of SARS-CoV-2 waves and vaccination timeline with time of sample collection is depicted in **Fig. S1**. Prior infection or breakthrough infection (BTI) were determined by a positive PCR or by Nucleocapsid (N) seroconversion or a two-fold increase in nucleocapsid IgG. The demographic and clinical characteristics of participants, at each timepoint, are summarized in **Table 1**. The study was approved by the University of Cape Town Human Research Ethics Committee (HREC 190/2020 and 291/2020), and written informed consent was obtained from all participants

### METHOD DETAILS

#### Isolation of PBMC

Blood was collected in heparin tubes and processed within 4 hours of collection. PBMC were isolated by density gradient sedimentation using Ficoll-Paque (Amersham Biosciences, Little Chalfont, UK) as per the manufacturer’s instructions. The PBMC were then cryopreserved in freezing media consisting of heat-inactivated fetal bovine serum (FBS, Thermofisher Scientific, Waltham, MA, USA) containing 10% dimethyl sulfoxide (DMSO) and stored in liquid nitrogen until use.

#### SARS-CoV-2 antigens

To measure SARS-CoV-2 spike-specific T-cell responses, we used custom mega pools of peptides. These peptides (15-mers overlapping by 10 amino acids) spanned the entire spike protein corresponding to the ancestral Wuhan sequence (GenBank: MN908947), Omicron B.1.1.529 (BA.1), XBB.1 and BA.2.86. The list of mutations for the Omicron sub-lineage compared to the ancestral Wuhan sequence is provided in **Table S1**. To measure SARS-CoV-2 nucleocapsid and membrane protein T-cell responses, we used commercial synthetic SARS-CoV-2 Pep-Tivator peptides (Miltenyi Biotec, Woking, UK), consisting of 15-mer sequences with 11 amino acids overlap, covering the complete sequence of the nucleocapsid (N, GenBank MN908947.3, Protein QHD43423.2) and membrane protein (M, GenBank MN908947.3, Protein QHD43419.1).

#### Cell stimulation and flow cytometry staining

Cryopreserved PBMC were thawed, washed, and rested for 4 hours in RPMI 1640 (Sigma-Aldrich, St Louis, MO, USA) supplemented with 10% heat inactivated FBS. After resting, cells were seeded in a 96-well V-bottom plate at ∼2 x10^6^ cells/well. Cells were stimulated with SARS-CoV-2 mega pools spanning the entire Spike (S) protein of the ancestral, Omicron BA.1, XBB.1 and BA.2.86 variants (1 µg/mL), as well the ancestral Membrane (M) and Nucleocapsid (N) proteins. All stimulations were performed in the presence of Brefeldin A (10 μg/mL, Sigma-Aldrich) and co-stimulatory antibodies against CD28 (clone 28.2) and CD49 (clone L25) (1 μg/mL each; BD Biosciences, San Jose, CA, USA). As a background control, PBMC were incubated with co-stimulatory antibodies, Brefeldin A and an equimolar amount of DMSO. After 16 hours of stimulation, cells were washed, stained with stained with LIVE/DEAD™ Fixable Near-IR Stain (Invitrogen, Carlsbad, CA, USA) and subsequently surface stained with the following antibodies: CD14 APC-Cy7 (HCD14, Biolegend, San Diego, CA, USA), CD19 APC-Cy7 (HIB19, Biolegend), CD4 PE-Cy7 (L200, BD Biosciences), CD8 BV510 (RPA-8, Biolegend), CD27 PE-Cy5 (1A4, Beckman Coulter, Brea, CA, USA) and CD45RA BV605 (HI100, Biolegend). Cells were then fixed and permeabilized using Cytofix/Cytoperm buffer (BD Biosciences) and stained with CD3 BV785 (OKT3), TNF-a FITC (Mab11) and IL-2 PE/Dazzle™ 594 (MQ1-17H12) from Biolegend and IFN-g Alexa 700 (B27, BD Biosciences). After intracellular cytokine staining, cells were washed and fixed in 1% Paraformaldehyde (ThermoFisher Scientific). Samples were acquired on a BD Fortessa using FACSDiva software and analysed using FlowJo (v10, FlowJo LLC, Ashland, OR, USA). Cells were gated on singlets, CD14^-^CD19^-^, live CD3+ T cells. Results are expressed as the frequency of total CD4+ or CD8+ T cells expressing IFN-γ, TNF-α or IL-2. The gating strategy is presented is **Fig. S4**. Due to high TNF-α backgrounds, cells producing TNF-α alone were excluded from the analysis. All data are presented after background subtraction (from the frequency of cytokine produced in unstimulated cells). To define the memory phenotype of SARS-CoV-2-specific T cells, a cut-off of 30 events was used.

### QUANTIFICATION AND STATISTICAL ANALYSIS

Statistical tests were performed using Prism (v10.3.1; GraphPad Software Inc, San Diego, CA, USA). Non-parametric tests were used for all comparisons. The Friedman test with Dunn’s correction was used for multiple group comparison, the Spearman rank test for correlation, and the Wilcoxon matched pairs test for paired samples. A P value <0.05 was considered statistically significant. Details of statistical analyses performed for each experiment are described in the figure legends.

---

**Supplementary Figure S1.**
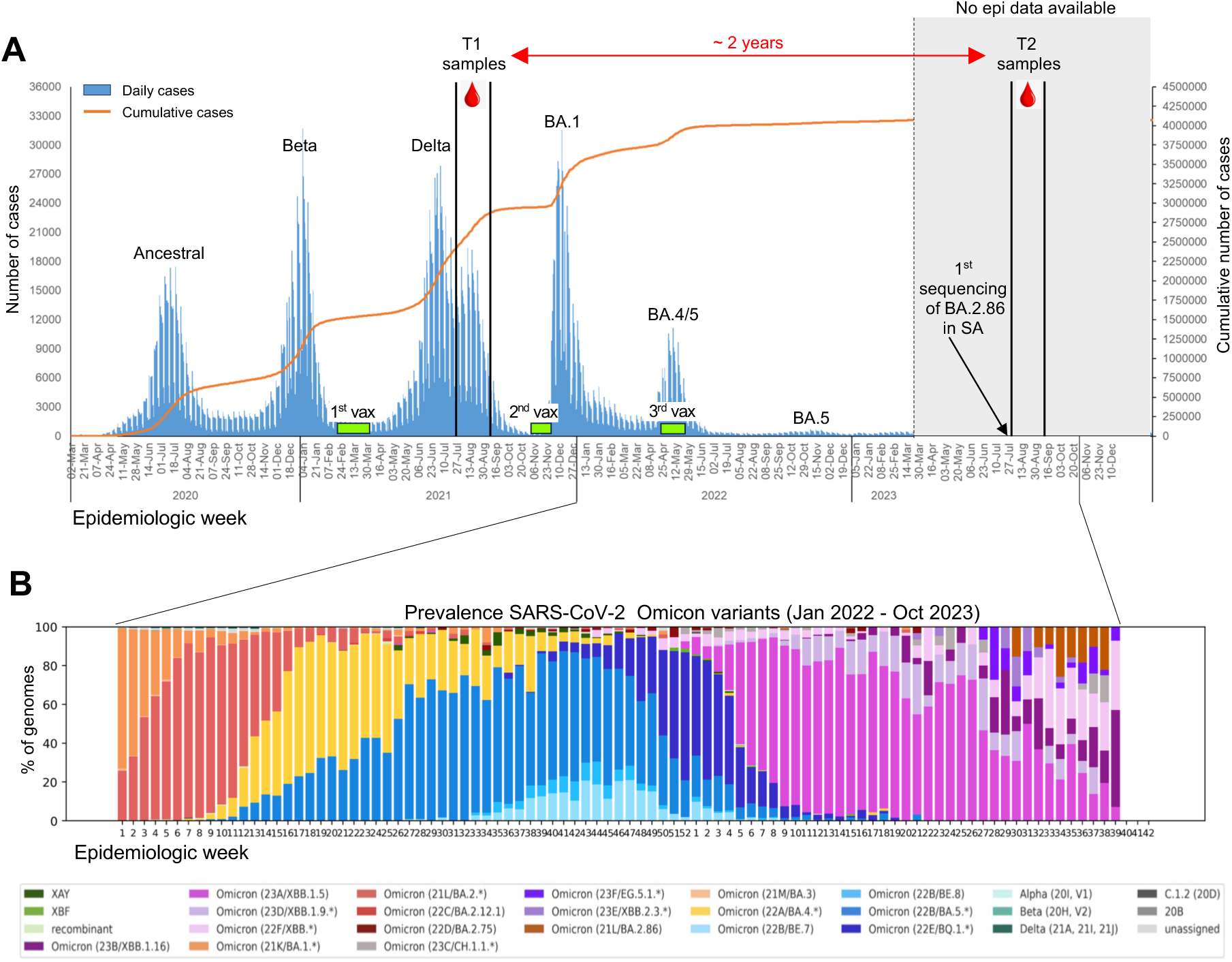
Landscape of SARS-CoV-2 waves and vaccination timeline in South Africa with time of sample collection. **(A)** T1 samples were collected between July and September 2021 during the second phase of the Delta wave and after the first vaccination campaign. T2 samples were collected approximately 2 years after T1 between July and September 2023. In the studied cohort, most of the participants (92.5%) were vaccinated with the Ad26.COV2.S vaccine. Two participants received a heterologous vaccination regimen (Ad26.COV2.S and BNT162b2) and one participant received 3 doses of the BNT162b2 vaccine. **(B)** Prevalence of Omicron sub-lineages between January 2022 and October 2023 based on 19,900 South African SARS-CoV-2 sequences from GISAID (www.gisaid.org). Epidemiologic and genomic surveillance data were obtained from the Network for Genomic Surveillance in South Africa (NGS-SA), National Institute for Communicable Diseases (NICD) of the National Health Laboratory (NHLS). https://www.nicd.ac.za/diseases-a-z-index/disease-index-covid-19/surveillance-reports/.

**Supplementary Figure S2.**
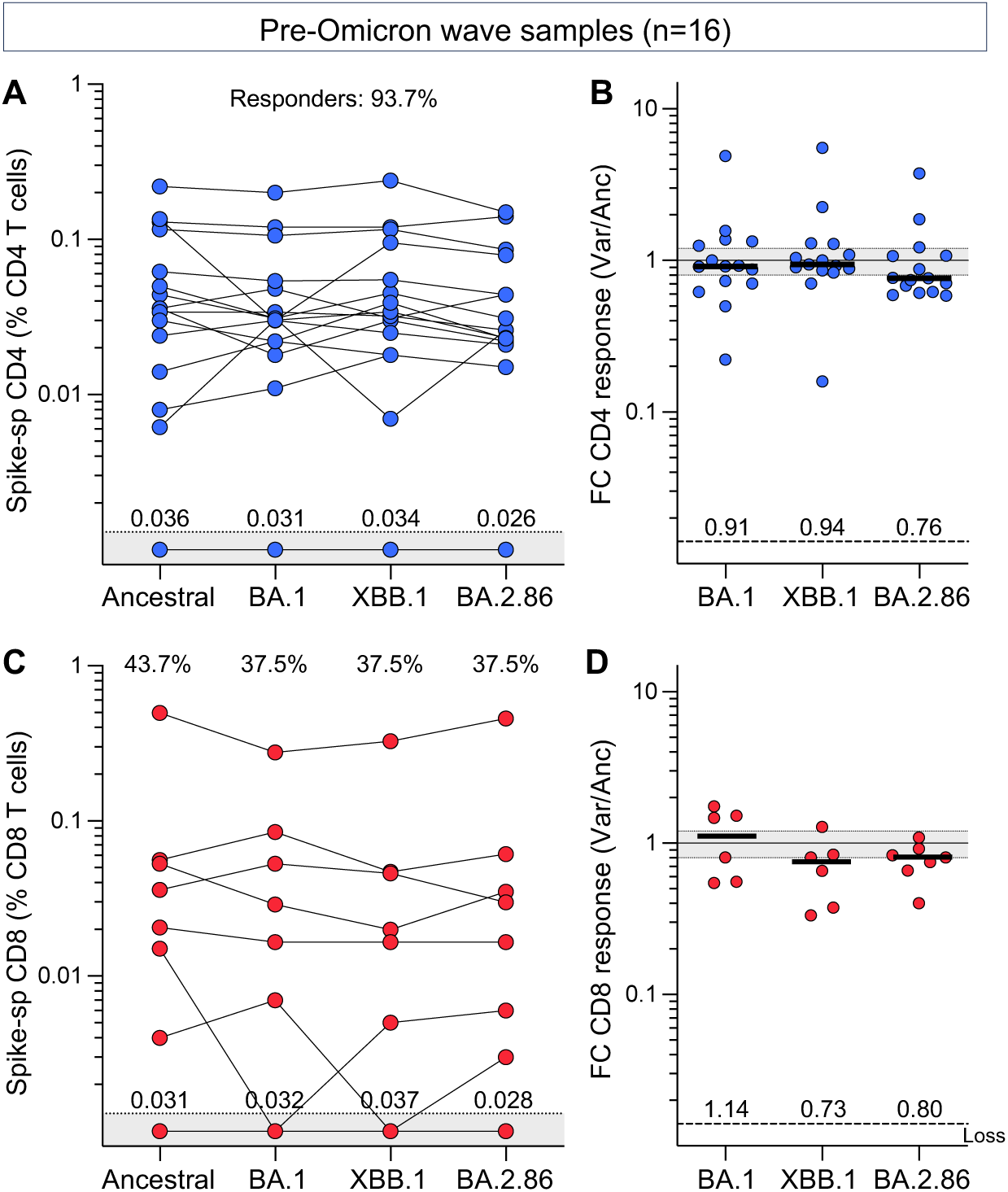
CD4+ and CD8+ T cell response to the SARS-CoV-2 ancestral, BA.1, XBB.1 or BA.2.86 spike in pre-Omicron participants. **(A and C)** Frequency of spike-specific CD4+ T cells (A) and spike-specific CD8+ T cells (C) producing any of the measured cytokines (IFN-γ, IL-2 or TNF-α) in 16 participants who were sampled prior to the emergence of the Omicron wave (July to Sept 2021). The proportion of responders is indicated at the top of the graphs. Median frequencies of spike-specific T cells in responders are indicated at the bottom of the graphs. **(B and D)** Fold change in the frequency of spike-specific CD4+ T cells (B) and spike-specific CD8+ T cells (D) between ancestral SARS-CoV-2 and variants. Median fold change in responders is represented by a line and indicated at the bottom of the graphs. Non-cross-reactive responses (‘loss’) are depicted at the bottom. No significant differences were observed between variants using a Friedman test with Dunn’s multiple comparisons post-test.

**Supplementary Figure S3.**
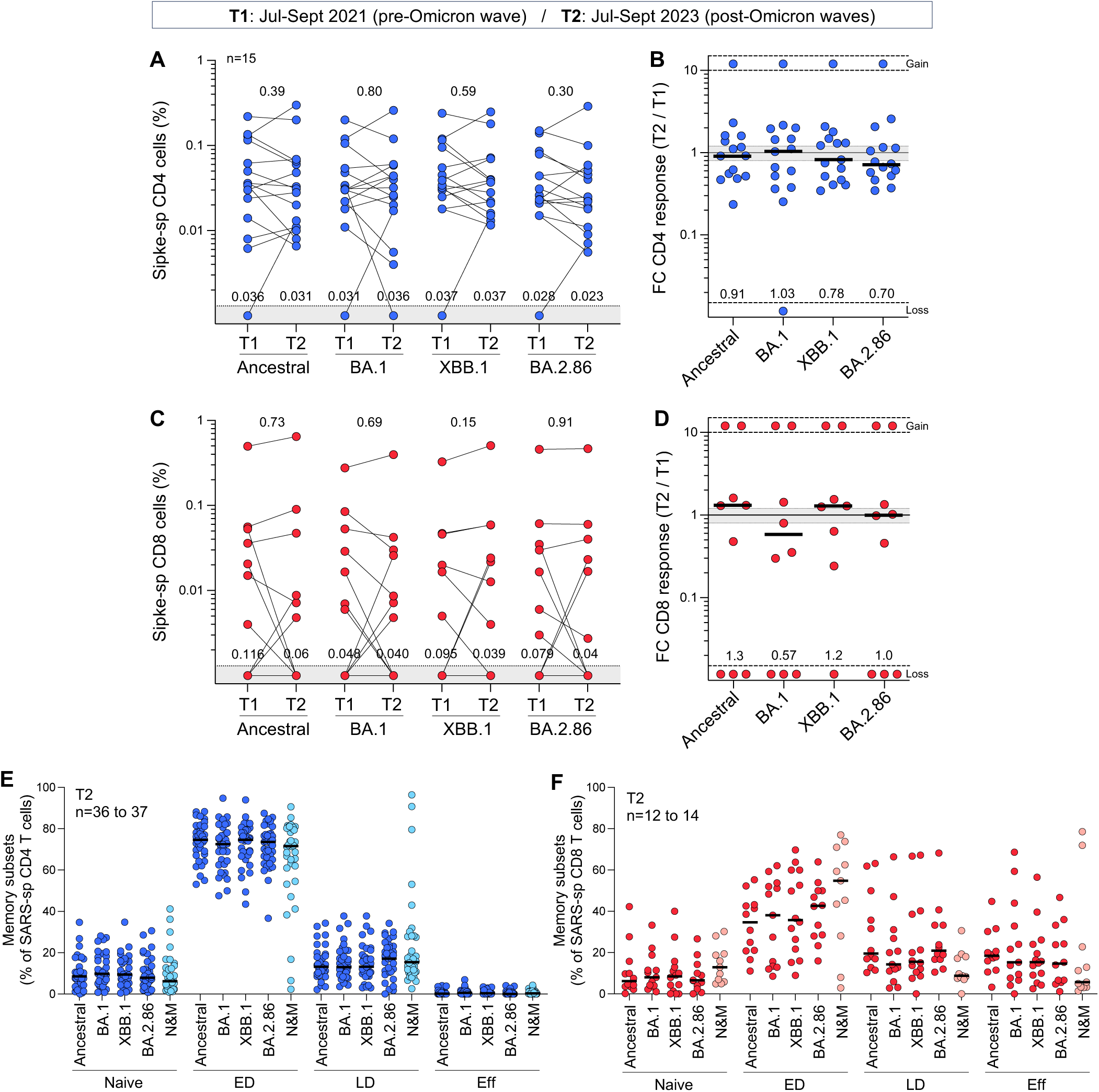
Longitudinal assessment of the maintenance and memory profile of SARS-CoV-2 ancestral spike-T cell response over 2 years. **(A and C)** Frequency of ancestral, BA.1, XBB.1 and BA.2.86 spike-specific CD4+ (A) and CD8+ T cells (C) in paired samples (n=15). T1 and T2 samples were collected between July and September 2021 and July and September 2023, respectively. Medians of responders are indicated at the bottom of the graphs. Statistical comparisons were assessed using a Wilcoxon matched-pairs signed rank test. **(B and D)** Fold change in the frequency of SARS-CoV-2 spike-specific CD4+ (B) and CD8+ T cells (D) between T2 and T1. Bars represent medians, and median fold change is indicated at the bottom of each graph. **(E-F)** Comparison of the memory profile of ancestral, BA.1, XBB.1, BA.2.86 spike-specific and N&M-specific CD4+ T cells (E) and CD8+ T cells (F) at T2. To define the memory phenotype of spike-specific T cells, a cut-off of 30 events was used.

**Supplementary Figure S4.**
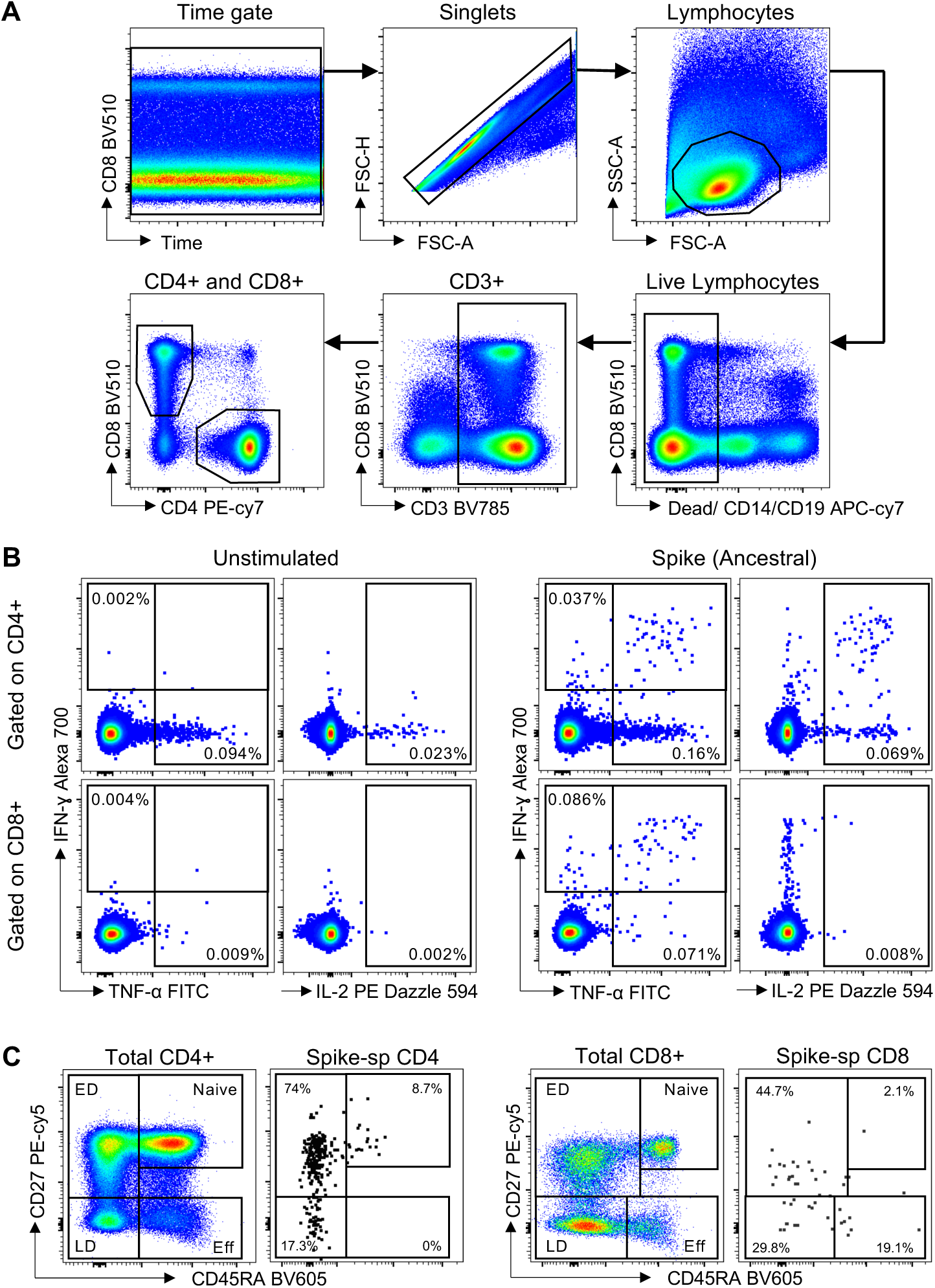
Gating strategy. **(A)** Nested gating strategy to identify CD4+ and CD8+ T cell populations. **(B)** Expression of IFN-γ, IL-2 and TNF-α in CD4+ and CD8+ T cells upon stimulation with spike peptide pool. **(C)** Identification of memory subsets in the total and SARS-CoV-2-specific CD4+ T cells (left panel) and CD8+ T cells (right panel) based on the expression of CD27 and CD45RA. Naïve: CD45RA+CD27+, Early differentiated (ED): CD45RA-CD27+, Late differentiated (ED): CD45RA-CD27-, and Effector (Eff): CD45RA+CD27-.

**Supplementary Table S1:** List of spike amino acid mutations in BA.1, XBB.1 and BA.2.86 as compared to Wuhan-1. “-“: deletion.

| Position<br>aa | BA.1 | XBB.1 | BA.2.86 | Position<br>aa | BA.1 | XBB.1 | BA.2.86 |
| --- | --- | --- | --- | --- | --- | --- | --- |
| 16 |  |  | MPLFV | 376 |  | A | A |
| 19 |  | I | I | 403 |  |  | K |
| 21 |  |  | T | 405 |  | N | N |
| 24 |  | - | - | 408 |  | S | S |
| 25 |  | - | - | 417 | N | N | N |
| 26 |  | - | - | 440 | K | K | K |
| 27 |  | S | S | 445 |  | P | H |
| 50 |  |  | L | 446 | S | S | S |
| 67 | V |  |  | 450 |  |  | D |
| 69 | - |  | - | 452 |  |  | W |
| 70 | - |  | - | 460 |  | K | K |
| 83 |  | A |  | 477 | N | N | N |
| 95 | I |  |  | 478 | K | K | K |
| 127 |  |  | F | 481 |  |  | K |
| 142 | D | D | D | 483 |  |  | - |
| 143 | - |  |  | 484 | A | A | K |
| 144 | - | Q | - | 486 |  | S | P |
| 145 | - |  |  | 490 |  | S |  |
| 146 |  | - |  | 493 | R |  |  |
| 152 | W |  |  | 496 | S |  |  |
| 157 |  |  | S | 498 | R | R | R |
| 158 |  |  | G | 501 | Y | Y | Y |
| 183 |  | E |  | 505 | H | H | H |
| 211 | - |  | - | 547 | K |  |  |
| 212 | I |  | I | 554 |  |  | K |
| 213 |  | E | G | 570 |  |  | V |
| 214 | EPE |  |  | 614 | G | G | G |
| 216 |  |  | F | 621 |  |  | S |
| 245 |  |  | N | 655 | Y | Y | Y |
| 252 |  | V |  | 670 |  |  | V |
| 264 |  |  | D | 679 | K | K | K |
| 332 |  |  | V | 681 | H | H | R |
| 339 | D | H | H | 764 | K | K | K |
| 346 |  | T |  | 796 | Y | Y | Y |
| 356 |  |  | T | 856 | K |  |  |
| 368 |  | I |  | 939 |  |  | F |
| 371 | L | F | F | 954 | H | H | H |
| 373 | P | P | P | 969 | K | K | K |
| 375 | F | F | F | 981 | F |  |  |
|  |  |  |  | 1143 |  |  | L |

